# Lung Ultrasound Findings in Patients Hospitalized with Covid-19

**DOI:** 10.1101/2020.06.25.20140392

**Authors:** Andre Kumar, Sukyung Chung, Youyou Duanmu, Sally Graglia, Farhan Lalani, Kavita Gandhi, Viveta Lobo, Trevor Jensen, Yingjie Weng, Jeffrey Nahn, John Kugler

## Abstract

**Introduction:** Point-of-care ultrasound (POCUS) has the potential to transform healthcare delivery in the era of COVID-19 with its diagnostic and therapeutic expediency. It can be performed by clinicians already at the bedside, which permits an immediate and augmented assessment of a patient. Although lung ultrasound can be used to accurately diagnose a variety of disease states such as pneumothorax, pleural effusions, pneumonia and interstitial lung disease2, there are limited reports on the sonographic manifestations of COVID-19. There is an urgent need to identify alternative diagnostic modalities that can be immediately employed at the bedside of COVID-19 patients.

**Methods:** This study was conducted at two medical centers in the United States from 3/21/2020-6/01/2020. Any adult who was hospitalized with COVID-19 (based on symptomatology and a confirmatory RT-PCR for SARS-CoV-2) and received a pulmonary POCUS examination was included. Providers were instructed to use a 12-zone scanning protocol for pulmonary views and save 6 second clips of each lung zone. This study utilized several POCUS devices, including Butterfly IQ, Vave, Lumify, and Sonosite. The collected images were interpreted by the study researchers based on a consensus document developed by the study authors and previously accepted definitions of lung POCUS findings.

**Results:** A total of 22 eligible patients who received 36 lung scans were included in our study. Eleven (50%) patients experienced clinical deterioration (defined as either ICU admission, invasive mechanical ventilation, or death within 28 days from the initial symptom onset). Among the 36 lung scans collected, only 3 (8%) were classified as normal. The remaining scans had the following abnormalities: presence of B-lines (n=32, 89%), consolidations (n=20, 56%), pleural thickening (n=17, 47%), and pleural effusion (n=4, 11%). Out of 20 scans with consolidations, 14 (70%) were subpleural and 5 (25%) were translobar. A-lines were present in 26 (72%) of patients, although they were only observed in the majority of the collected lung zones in 5 (14%) of patients. Ultrasound findings were stratified by time from symptom onset to the scan based on the following time periods: early (0-6 days), middle (7-13 days), and late (14-28 days). B-lines appeared early after symptom onset and persisted well into the late disease course. In contrast, pleural thickening increased in frequency over time (early: 25%, middle: 47%, late: 67%). Subpleural consolidations also appeared in higher frequency later in the disease course (early: 13%, middle 42%, late: 56%).

**Discussion:** certain lung ultrasound findings may be common in Covid-19, while others may appear later in the disease course or only occur in patients who experience clinical deterioration. Future efforts should investigate the predictive utility of consolidations, pleural thickening and B-lines for clinical deterioration and compare them to traditional radiological studies such as X-rays or CTs.

## Introduction

Point-of-care ultrasound (POCUS) has the potential to transform healthcare delivery in the era of COVID-19 with its diagnostic and therapeutic expediency.^1^ It can be performed by clinicians already at the bedside, which permits an immediate and augmented assessment of a patient.^2^ POCUS devices, particularly handheld devices, are often cheaper than traditional radiological equipment such as X-ray or computerized tomography (CT) machines, which makes POCUS ideal for surge scenarios and other resource-limited settings. Since providers using POCUS are concomitantly at the bedside assessing patients, POCUS may reduce personal protective equipment usage by radiological technicians or the need to decontaminate larger radiological equipment.

Although lung ultrasound can be used to accurately diagnose a variety of disease states such as pneumothorax, pleural effusions, pneumonia and interstitial lung disease^2^, there are limited reports on the sonographic manifestations of COVID-19. There is an urgent need to identify alternative diagnostic modalities that can be immediately employed at the bedside of COVID-19 patients. In this report, we characterize lung ultrasound findings of patients admitted to our hospitals with COVID-19 and stratify these findings by time, location, and illness severity.

## Methods

This study was conducted at two medical centers in the United States from 3/21/2020-6/01/2020. Any adult who was hospitalized with COVID-19 (based on symptomatology^3^ and a confirmatory RT-PCR for SARS-CoV-2) and received a pulmonary POCUS examination was included. Patients who did not meet these criteria were excluded. Our Institutional Review Board approved this study.

Provider discretion determined whether to perform an initial or follow-up POCUS examination for each patient. Providers were instructed to use a 12-zone scanning protocol for pulmonary views (Figure 1) and save 6 second clips of each lung zone.^4^ This study utilized several POCUS devices, including Butterfly IQ™, Vave™, Lumify™, and Sonosite™. The collected images were interpreted by the study researchers based on a consensus document developed by the study authors and previously accepted definitions of lung POCUS findings.^4-7^

**Figure 1.**
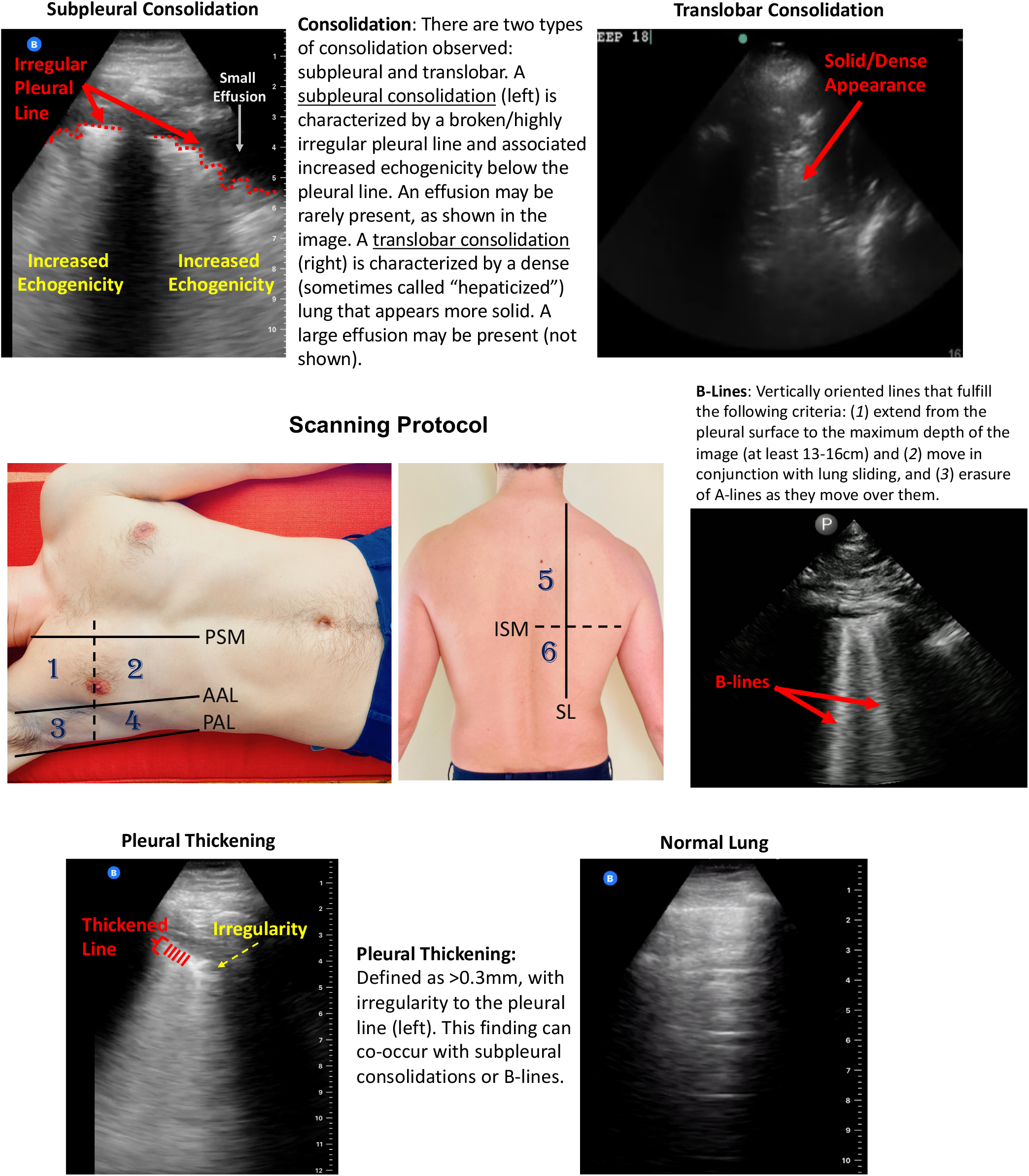
Scanning Protocol and Lung Ultrasound Findings in COVID-19 Patients. This study utilized a 12-zone protocol.^4^ On each hemithorax, there are 6 zones. The exam begins on the patient’s right side. Zones 1-2 (anterior zones) are between the parasternal margin (PSM) and the anterior axillary line (AAL) and are best obtained in the mid-clavicular line. Zones 3-4 (lateral zones) are between the anterior axillary line (AAL) and posterior axillary line (PAL) and are best obtained in the mid-axillary line. The nipple line serves as a bisecting area between these zones. Zones 5-6 (posterior zones) are medial to the scapular line (SL) and are bisected by the inferior scapular margin (ISM). The zone areas are repeated on the contralateral hemithorax (starting with zone 7). This figure contains an overview of the observed ultrasound findings based on previously described terminology.^4,6,7^

## Results

A total of 22 eligible patients who received 36 lung scans were included in our study (Table 1). Eleven (50%) patients experienced clinical deterioration (defined as either ICU admission, invasive mechanical ventilation, or death within 28 days from the initial symptom onset).

**Table 1.** Ultrasound Findings Among Patients Hospitalized with COVID-19. Patients were further stratified by clinical deterioration (defined as any occurrence of ICU admission, invasive mechanical ventilation, or death within 28 days from initial symptoms onset) Proportions are based on the number of scans for a given lung zone. Median and IQR are displayed where appropriate. BMI, Body Mass Index; IQR, interquartile range.

|  | All Patients | Clinical Deterioration | No Clinical Deterioration |
| --- | --- | --- | --- |
| **Demographics** |  |  |  |
| Number of Patients | 22 | 11 | 11 |
| Number of Scans Performed | 36 | 16 | 20 |
| Age (Years) [IQR] | 47 [33-72] | 43 [32-69] | 55 [33-87] |
| Body Mass Index, [IQR] | 29 [27-35] | 33 [27-36] | 28 [26-35] |
| Male (%) | 12 (55%) | 8 (72%) | 6 (55%) |
| **Clinical Characteristics** |  |  |  |
| Days from Symptom Onset to Scan [IQR] | 10 [6-13] | 12 [7-23] | 9 [6-12] |
| Admitted to ICU |  | 11 | 0 |
| Received Mechanical Ventilation |  | 6 | 0 |
| Death |  | 1 | 0 |
| **POCUS Findings** |  |  |  |
| Anterior Lung Zone Scans | 33 | 15 | 18 |
| Lateral Lung Zone Scans | 31 | 14 | 17 |
| Posterior Lung Zone Scans | 18 | 7 | 11 |
| Normal Scan (%) | 3 (8%) | 0 (0%) | 3 (15%) |
| A-lines Observed in $\geq 50\%$ of Zones (%) | 5 (14%) | 1 (6%) | 4 (20%) |
| B-lines Observed (%) | 32 (89%) | 16 (100%) | 16 (80%) |
| $\geq 3$ B-Lines in any Zone | 32 (89%) | 14 (88%) | 18 (90%) |
| Bilateral B-lines | 19 (53%) | 10 (63%) | 9 (45%) |
| Anterior Zone B-Lines | 23 (70%) | 13 (87%) | 10 (56%) |
| Lateral Zone B-Lines | 23 (74%) | 9 (64%) | 14 (82%) |
| Posterior Zone B-Lines | 12 (67%) | 5 (71%) | 7 (63.6%) |
| Zones per Scan with B-Lines [IQR] | 4 [1-8] | 6 [3-8] | 3.5 [1-8] |
| Consolidation (%) | 20 (56%) | 11 (69%) | 9 (45%) |
| Translobar Consolidation | 5 (25%) | 3 (19%) | 2 (10%) |
| Subpleural consolidation | 14 (70%) | 7 (44%) | 7 (35%) |
| Bilateral Consolidation | 11 (31%) | 8 (50%) | 3 (15%) |
| Anterior Consolidation | 9 (27%) | 7 (47%) | 2 (11%) |
| Lateral Consolidation | 15 (48%) | 10 (71%) | 5 (29%) |
| Posterior Consolidation | 9 (50%) | 3 (43%) | 6 (55%) |
| Pleural Effusion (%) | 4 (11%) | 4 (25%) | 0 (0%) |
| Bilateral Pleural Effusions | 3 (8%) | 3 (19%) | 0 (0%) |
| Pleural Thickening (%) | 17 (47%) | 11 (69%) | 6 (30%) |

Among the 36 lung scans collected, only 3 (8%) were classified as normal (Table 1). The remaining scans had the following abnormalities: presence of B-lines (n=32, 89%), consolidations (n=20, 56%), pleural thickening (n=17, 47%), and pleural effusion (n=4, 11%). Out of 20 scans with consolidations, 14 (70%) were subpleural and 5 (25%) were translobar. A-lines were present in 26 (72%) of patients, although they were only observed in the majority of the collected lung zones in 5 (14%) of patients. Table 1 also displays the above findings by location.

Scans from patients who experienced clinical deterioration demonstrated higher percentages of B-lines (100% vs. 80%), consolidations (69% vs. 45%), pleural thickening (69% vs. 30%) and effusions (25% vs. 0%). Additionally, scans from patients with clinical deterioration had higher percentages of bilateral B-lines (63% vs. 45%) and bilateral consolidation (50% vs. 15%).

Ultrasound findings were stratified by time from symptom onset to the scan based on the following time periods: early (0-6 days), middle (7-13 days), and late (14-28 days). B-lines appeared early after symptom onset and persisted well into the late disease course (Figure 2). In contrast, pleural thickening increased in frequency over time (early: 25%, middle: 47%, late: 67%). Subpleural consolidations also appeared in higher frequency later in the disease course (early: 13%, middle 42%, late: 56%). A-lines were observed throughout the time course (early: 63%, middle 74%, late 78%; Figure 2), although A-lines often occurred in a patchy distribution.

**Figure 2.**
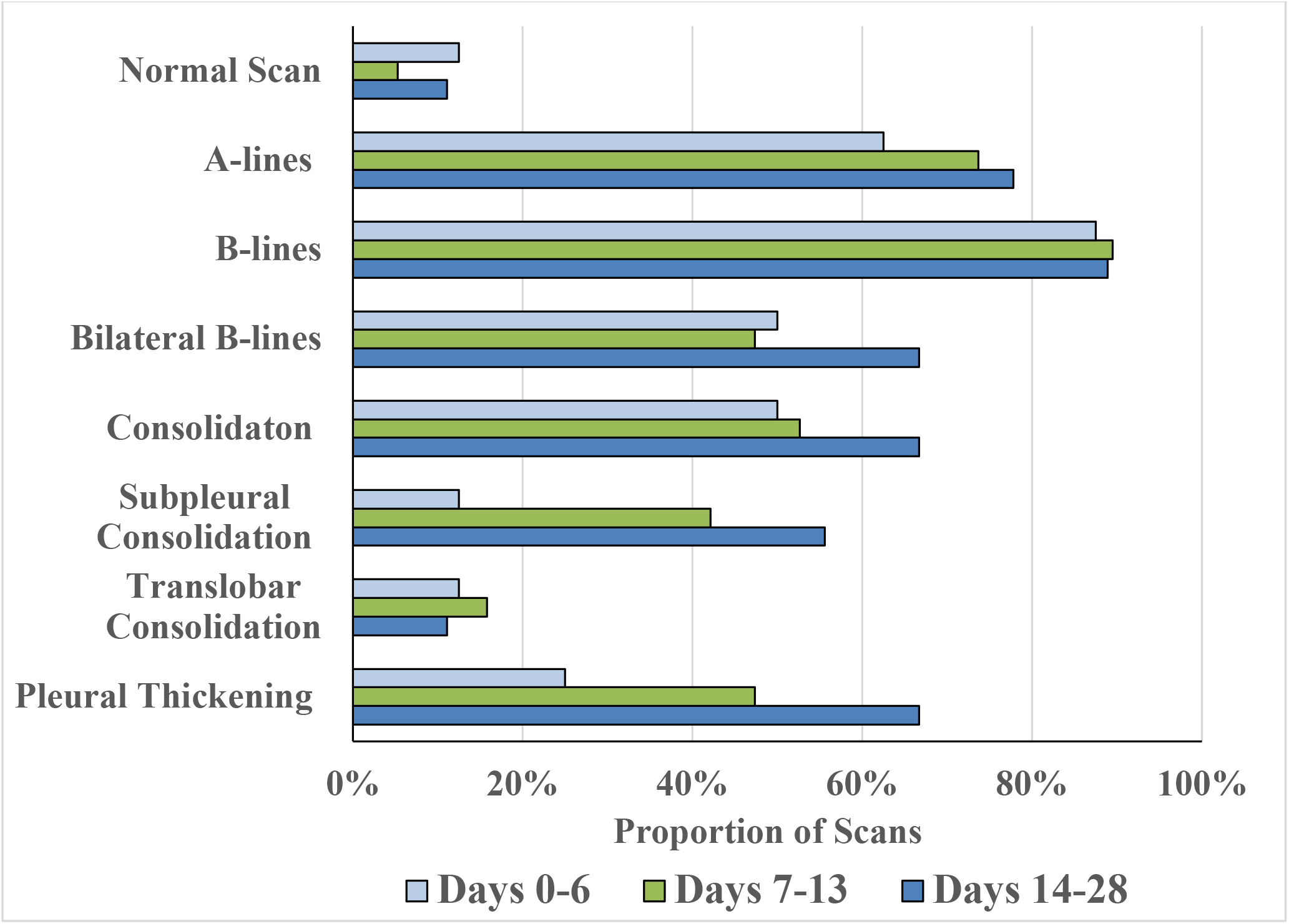
Distribution of Lung Ultrasound Findings by Time. Lung findings were stratified by days from symptom onset to the ultrasound scan in the following manner: early (0-6 days), middle (7-13 days), and late (14-28 days). The portion of scans that contained the above findings are displayed on the horizontal axis.

## Discussion

In this report, we characterize lung ultrasound findings for patients admitted to the hospital with COVID-19, and further stratify these findings by patients who experienced clinical deterioration. Common findings included B-lines, consolidation (including subpleural consolidations), and pleural thickening. Effusions were rare, which is consistent with CT studies from COVID-19 patients.^5^ Notably, B-lines were common for both patients who did and did not experience clinical deterioration. They frequently appeared in all lung zones, and they persisted throughout the 28 day scanning period. In contrast, subpleural consolidation and pleural thickening appeared later in the disease course, and they were more common in patients who experienced clinical deterioration. Importantly, bilateral involvement of several findings (B-lines or consolidation) was more commonly encountered in the patients who experienced clinical deterioration. Certain patient conditions, such as intubation or patient mobility, affected the provider from acquiring all 12 zones, particularly the posterior zones.

In conclusion, certain lung ultrasound findings may be common in Covid-19, while others may appear later in the disease course or only occur in patients who experience clinical deterioration. Future efforts should investigate the predictive utility of consolidations, pleural thickening and B-lines for clinical deterioration and compare them to traditional radiological studies such as X-rays or CTs.

## Data Availability

All summary data are available to individuals who request it. We cannot share data that contains PHI.

